# Genetic inhibition of interleukin-6 receptor signaling and Covid-19

**DOI:** 10.1101/2020.07.17.20155242

**Authors:** Jonas Bovijn, Cecilia M Lindgren, Michael V Holmes

## Abstract

There are few effective therapeutic options for the treatment of severe acute respiratory syndrome coronavirus 2 (SARS-CoV-2) infection. Early evidence has suggested that IL-6R blockers may confer benefit, particularly in severe coronavirus disease 2019 (Covid-19).

We leveraged large-scale human genetic data to investigate whether IL6-R blockade may confer therapeutic benefit in Covid-19. A genetic instrument consisting of seven genetic variants in or close to *IL6R* was recently shown to be linked to altered levels of c-reactive protein (CRP), fibrinogen, circulating IL-6 and soluble IL-6R, concordant to known effects of pharmacological IL- 6R blockade. We investigated the effect of these *IL6R* variants on risk of hospitalization for Covid- 19 and other SARS-CoV-2-related outcomes using data from The Covid-19 Host Genetics Initiative.

The *IL6R* variants were strongly associated with serum CRP levels in UK Biobank. Meta-analysis of scaled estimates revealed a lower risk of rheumatoid arthritis (OR 0.93 per 0.1 SD lower CRP, 95% CI, 0.90-0.96, *P* = 9.5 × 10^−7^), recapitulating this established indication for IL-6R blockers (e.g. tocilizumab and sarilumab). The IL-6R instrument was associated with lower risk of hospitalization for Covid-19 (OR 0.88 per 0.1 SD lower CRP, 95% CI, 0.78-0.99, *P* = 0.03). We found a consistent association when using a population-based control group (i.e. all non-cases; OR 0.91 per 0.1 SD lower CRP, 95% CI, 0.87-0.96, *P* = 4.9 × 10^−4^). Evaluation of further SARS-CoV-2-related outcomes suggested association with risk of SARS-CoV-2 infection, with no evidence of association with Covid-19 complicated by death or requiring respiratory support. We performed several sensitivity analyses to evaluate the robustness of our findings.

Our results serve as genetic evidence for the potential efficacy of IL-6R blockade in Covid-19. Ongoing large-scale RCTs of IL-6R blockers will be instrumental in identifying the settings, including stage of disease, in which these agents may be effective.

---

There are few effective therapeutic options for the treatment of severe acute respiratory syndrome coronavirus 2 (SARS-CoV-2) infection. Interleukin-6 receptor (IL-6R) blockade has been proposed as one potential therapeutic strategy and more than 40 clinical trials for anti-IL-6R antibodies (including tocilizumab and sarilumab) in the setting of SARS-CoV-2 infection are currently underway. Early evidence—from observational studies and open-label, uncontrolled trials—has suggested that IL-6R blockers may confer benefit, particularly in severe coronavirus disease 2019 (Covid-19).^1^

We leveraged large-scale human genetic data^2^ to investigate whether IL6-R blockade may confer therapeutic benefit in Covid-19. A genetic instrument consisting of seven genetic variants in or close to *IL6R* (pairwise r^2^ ≤ 0.1) was recently shown to be linked to altered levels of c-reactive protein (CRP), fibrinogen, circulating IL-6 and soluble IL-6R, concordant to known effects of pharmacological IL-6R blockade.^3^ We investigated the effect of these *IL6R* variants on risk of hospitalization for Covid-19 and other SARS-CoV-2-related outcomes using data from The Covid- 19 Host Genetics Initiative.^4^

The *IL6R* variants were strongly associated with serum CRP levels in UK Biobank (**Figure 1A**). Meta-analysis of scaled estimates revealed a lower risk of rheumatoid arthritis (OR 0.93 per 0.1 SD lower CRP, 95% CI, 0.90-0.96, *P* = 9.5 × 10^−7^; **Figure 1B**), recapitulating this established indication for IL-6R blockers (e.g. tocilizumab and sarilumab), and coronary heart disease (OR 0.96, 95% CI, 0.95-0.98, *P* = 2.0 × 10^−6^), which has previously been linked to genetic variation in *IL6R*.^5^ *The IL-6R instrument was associated with lower risk of hospitalization for Covid-19 (OR 0*.*88 per 0*.*1 SD lower CRP, 95% CI, 0*.*78-0*.*99, P* = 0.03; **Figure 1B**). We found a consistent association when using a population-based control group (i.e. all non-cases; OR 0.91 per 0.1 SD lower CRP, 95% CI, 0.87-0.96, *P* = 4.9 × 10^−4^; **Figure 1B**).

**Figure 1.**
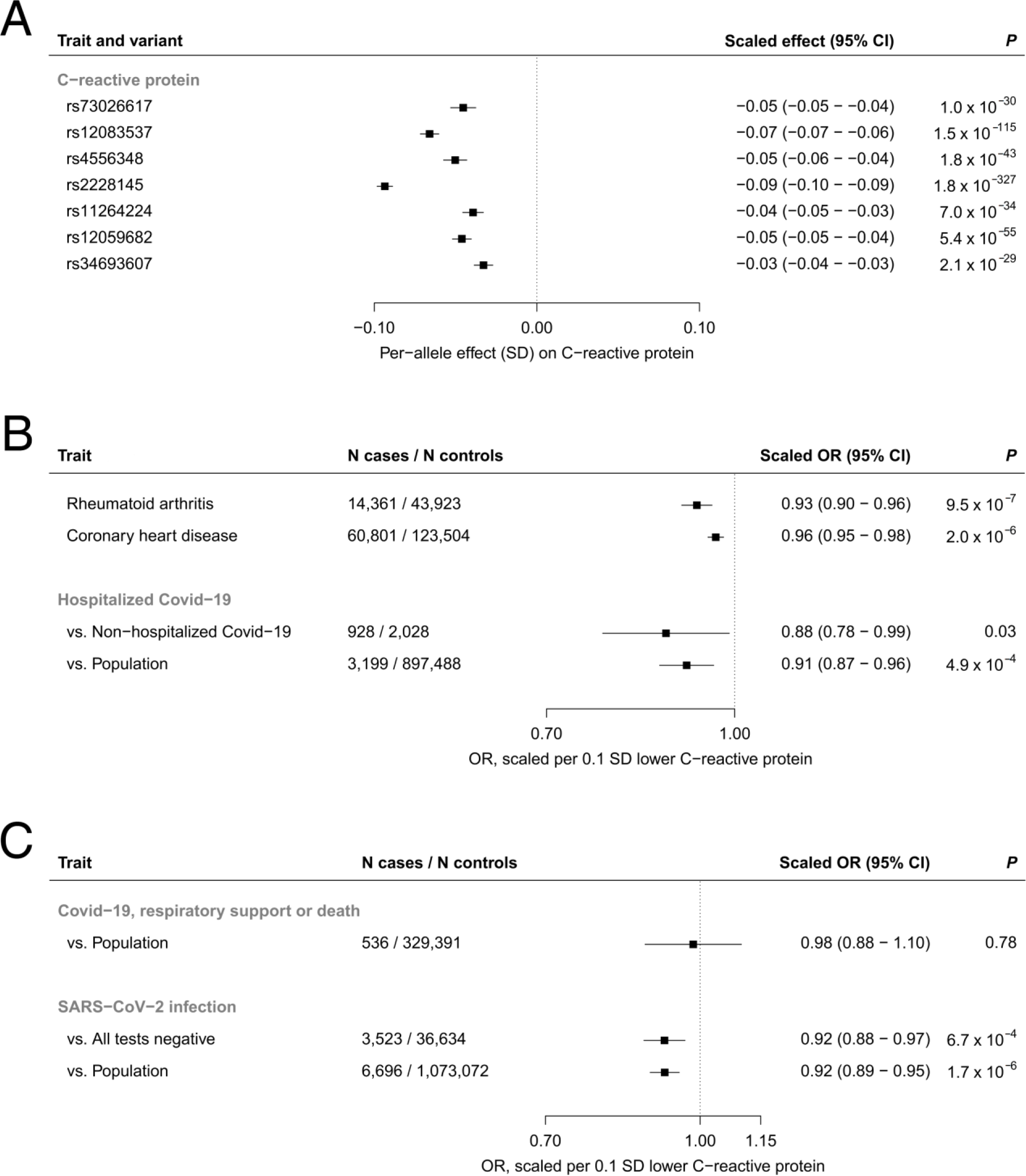
Association of CRP-lowering *IL6R* variants with SARS-CoV-2-related outcomes. Panel A shows the per-allele association of 7 independent CRP-lowering variants in or near *IL6R* with CRP in UK Biobank. Panel B shows the scaled association of the *IL6R* instrument with two positive controls (rheumatoid arthritis, given the established indication of IL-6R blockers, and coronary heart disease, which has been previously linked to variants in *IL6R*) and risk of hospitalization for confirmed SARS-CoV-2 infection. Risk of hospitalization was assessed using a control group of individuals with confirmed SARS-CoV-2 infection and no hospitalization at 21 days post-test, and a population-based control group of all non-cases. Panel C shows the scaled associations of the *IL6R* instrument with secondary outcomes (Covid-19 complicated by death or requiring respiratory support, and risk of SARS-CoV-2 infection, as defined by laboratory confirmed SARS-CoV-2 infection and/or a confirmed diagnosis of Covid-19). See Supplementary Appendix for definitions of outcomes used in The Covid-19 Host Genetics Initiative. OR, odds ratio; CI, confidence interval.

Evaluation of further SARS-CoV-2-related outcomes suggested association with risk of SARS-CoV-2 infection, with no evidence of association with Covid-19 complicated by death or requiring respiratory support (**Figure 1C**). We performed several sensitivity analyses to evaluate the robustness of our findings (see Supplementary Appendix for further details, including phenotype definitions and results).

Our findings show that *IL6R* variants mimicking therapeutic inhibition of IL-6R are associated with lower risk of being hospitalized for Covid-19, a phenotype that correlates with disease severity (e.g. requiring supplemental oxygen is a typical reason for hospitalization). This suggests that pharmacological IL-6R blockade may be expected to lead to reduced Covid-19 severity. We also found an association with lower risk of SARS-CoV-2 infection. Whilst the latter finding may suggest that IL-6R blockade lowers susceptibility to SARS-CoV-2 infection, these phenotypes may be biased by symptom severity (e.g. individuals with more severe symptoms may be more likely to present for testing, to be offered testing or to have a positive test). The lack of association with very severe Covid-19 requiring respiratory support or leading to death may bear relevance in the context of the recently announced failure of sarilumab (a recombinant human monoclonal antibody against IL-6R) in a phase III RCT in patients requiring mechanical ventilation.^6^ However, the genetic analysis with very severe Covid-19 was based on the fewest cases (536), which limits robust inference.

## Data Availability

All data included in this study are publicly available. All sources are stated in the manuscript.

## Acknowledgments

We express our gratitude to the studies and consortia (including the Neale lab, the Covid-19 Host Genetics Initiative, the CARDIoGRAMplusC4D consortium and Okada et al.), in particular their participants and their investigators, for providing access to summary statistics.

## Funding

J.B. is supported by funding from the Rhodes Trust, Clarendon Fund and the Medical Sciences Doctoral Training Centre, University of Oxford. C.M.L. is supported by the Li Ka Shing Foundation, WT-SSI/John Fell funds, and the National Institute for Health Research Oxford Biomedical Research Centre; M.V.H. works in a unit that receives funding from the UK Medical Research Council and is supported by a British Heart Foundation Intermediate Clinical Research Fellowship (FS/18/23/33512) and the National Institute for Health Research Oxford Biomedical Research Centre.

## Declaration of interests

J.B. has served as a consultant to the Bill and Melinda Gates Foundation Strategic Investment Fund. C.M.L. has collaborated with Novo Nordisk and Bayer in research, and in accordance with a university agreement, did not accept any personal payment. M.V.H. has collaborated with Boehringer Ingelheim in research, and in accordance with the policy of the Clinical Trial Service Unit and Epidemiological Studies Unit (University of Oxford), did not accept any personal payment.

## Contributors

J.B. conceived of and designed the study, analysed the data and wrote the first draft of the manuscript. All authors contributed to the interpretation of findings and writing of subsequent versions of the manuscript.

